# Liver Radioembolization: Predicting Lung Shunt Fraction with Contrast-Enhanced CT, Eliminating the Need for 99mTc-MAA

**DOI:** 10.1101/2024.12.21.24319495

**Authors:** Brahim Mehadji, Talia Marx, Adrianna Carter, Roger Goldman, Catherine Vu, Emilie Roncali

## Abstract

1.

Accurate estimation of the Lung Shunt Fraction (LSF) is a standard of care in yttrium-90 (^90^Y) radioembolization treatment planning to prevent excessive lung irradiation due to arterio-venous shunting in the liver. LSF is assessed using ^99m^Tc macroaggregated albumin (^99m^Tc-MAA) imaging, but this approach adds risk, complexity, and expense to the treatment planning. This study investigates the potential of Contrast-Enhanced Computed Tomography (CECT) as a non-invasive alternative for LSF estimation.

**Methods:** We developed a novel metric to estimate the LSF from differences between CECT phases, based on the premise that LSF results from tumor angiogenesis and arterio-venous shunting that create excessive local opacity in the CECT arterial phase. Hypervascular volumes were identified by subtracting the portal phase from the arterial phase and contouring the corresponding regions using an density threshold. The perfused volume corresponded to the lobe and to the volume injected with ^99m^Tc-MAA.

We conducted a retrospective analysis involving 30 liver cancer patients who underwent 4-phase liver CECT and ^99m^Tc-MAA imaging prior to ^90^Y radioembolization to study several markers based on the subtraction images. The patient cohort included several types of liver tumors (primary or metastatic), predominantly hepatocellular carcinoma. Correlations between the hypervascular-to-perfused volume ratio from CECT and LSF values from standard-of-care ^99m^Tc-MAA planar imaging were assessed.

**Results:** The strongest correlation was obtained between the hypervascular-to-perfused volume ratio from CECT and LSF values from ^99m^Tc-MAA planar gamma imaging (R²=0.95). The best correlation was demonstrated when the hypervascular volume was corrected by the hypoxic volume and then normalized by the perfused volume. In contrast, the tumor volume did not show a strong correlation with LSF (R²=0.38). Moreover, the predictive power of the hypervascular-to-perfused volume ratio on LSF demonstrated a 1-sigma uncertainty of 3%.

**Conclusion:** CECT-based LSF estimation shows promise as a non-invasive alternative to ^99m^Tc-MAA imaging in ^90^Y treatment planning. Further validation with prospective studies on larger cohorts is necessary to confirm the accuracy and safety of this approach.

## 2 Introduction

Radioembolization is widely used for early and advanced liver cancer treatment, including unresectable hepatocellular carcinoma (HCC) and metastatic liver tumors. Benefits include improved quality of life, increased time to progression as comparted to chemoembolization^1^, safety in treatment of patients with bland or malignant portal vein thrombosis^2^, and downstaging for liver transplant^3–9^. Radioembolization has been adopted as a primary treatment for resectable and unresectable HCC^10^ and represents more than 10,000 interventions in the U.S. annually^11^.

Radioembolization consists of administering radioactive yttrium-90 (^90^Y) glass or resin microspheres (30 µm average diameter) through a catheter positioned in the hepatic arteries with arterial access typically from the common femoral or radial artery^12,13^. Transarterial injection of microspheres performed in interventional radiology (IR) requires multiple imaging modalities and radiology subspecialties. Planning for Y90 transarterial radioembolization typically requires an invasive procedure that includes vascular mapping and a pre-treatment Tc-99m macro-aggregated albumin (MAA), performed in a short time interval 0-14 days prior to treatment. ^99m^Tc-MAA particles are injected in the liver arterial blood flow and their distribution is imaged with planar gamma imaging or single photon computed tomography (SPECT) to evaluate the lung shunt fraction (LSF). The LSF predicts the fraction of injected ^90^Y microspheres that may reach the lungs through abnormal vasculature and cause lung toxicity^14^. This exam adds a significant burden to the treatment planning, requiring administration of a diagnostic radiopharmaceutical and nuclear medicine imaging study which extends the planning time by several hours.

Clinical radioembolization practice has dramatically evolved in the last 10 years, with much greater doses delivered in a highly targeted manner as done in radiation segmentectomy^8,15,16^. There is a growing interest in bypassing the workup for segmentectomy but long-term impact on patient care is not fully known^17–19^. Predicting the ^90^Y dose distribution before treatment is considered helpful to ensure the safety of the treatment and adequacy of the injected dose^20,21^. Developing strategies promoting streamlined treatment planning is thus becoming important.

A potential solution for simplifying the planning LSF estimation is to use pre-treatment imaging such as Contrast-Enhanced Computed Tomography (CECT) or magnetic resonance imaging (MRI) which are routinely performed to visualize and measure important characteristics such tumor volume, hepatic arterial and venous structures. Both these modalities have also shown a correlation between liver perfusion and patient outcome^22–26^. Two main steps are necessary to re-design treatment planning without a visit to the IR suite. The first aspect is to develop predictive dosimetry for ^90^Y radioembolization, which we recently demonstrated with a CECT-based approach^27^. The second aspect is to develop a method to predict the LSF without ^99m^Tc-MAA SPECT, which is the focus of this paper.

Liver CECT often consists of a 4-phase CT scan with an acquisition before contrast injection (baseline), a scan shortly post-injection (p.i.) to capture the liver arterial blood flow (∼30-40 sec p.i.), a portal phase scan highlighting the venous and portal blood flows (∼70-90 sec p.i.), and a late equilibrium phase (∼60-180 sec p.i.)^27^. Similarly to our previous work manipulating CECT images to predict the ^90^Y dose distribution, we propose and develop a novel approach to calculate a metric indicative of the LSF solely based on differences between CECT phases. Tumor volume alone does not reliably predict LSF due to tumor variability, including differences in vascular supply, structure and the presence of hypoxic regions within the tumor^28^. Our approach is based on the hypothesis that LSF is related to tumor angiogenesis that leads to arterio-venous shunting^29^. Thus, voxels within a tumor with high density in arterial phase likely indicate the presence of arterio-venous shunting. This LSF contributor is determined by subtracting the portal phase from the arterial phase, identifying regions with null or positive values, and calculating their volume. Areas with low density in both phases, presumed to represent hypoxic regions, are excluded from this volume. We analyzed various markers derived from the subtracted images and demonstrated a strong correlation with the reference LSF, obtained from 30 clinical planar gamma images taken pre-radioembolization.

## 3 Materials and Methods

### 3.1 Patient population

In this retrospective study, 30 patients with documented hepatic lesions who had a 4-phase liver CECT scan within 1 month prior to their workup were selected. Twenty-four patients presented with hepatocarcinoma (HCC), and two patients had cholangiocarcinoma. The remaining four patients presented with metastatic lesions from neuroendocrine tumors, gastrinoma, colorectal adenocarcinoma, and lymphocyte carcinoma, respectively.

All patients underwent standard-of-care radioembolization workup which included a ^99m^Tc-MAA injection. A single injection of ^99m^Tc-MAA was administered via the right hepatic artery in 24 patients and via the left hepatic artery in five patients. One patient received the injection in a segmental artery of the right lobe.

### 3.2 Standard-of-care LSF estimation with ^99m^Tc-MAA imaging

Planar gamma imaging of the lungs and liver was conducted within two hours of the ^99m^Tc-MAA injection, on a GE Hawkeye (GE Healthcare, Chicago, IL) or a Philips BXCT (Philips, Amsterdam, Netherlands) system. Planar imaging and SPECT/CT both constitute clinical standards for estimating lung shunt fraction. Planar imaging is more widely used than SPECT/CT for practical reasons^30^. We retrospectively collected LSF values calculated by board-certified radiologists following the European Association of Nuclear Medicine (EANM) guidelines^31^ and below defined workflow. Regions of interest (ROIs) encompassing the liver and lungs as well as background ROIs were placed on anterior and posterior images. The number of counts in each region, *Norgan,raw,ant/post* was corrected by the background *Nbckg* to obtain the net organ count *Norgan,ant/post* using the following equations^32^:

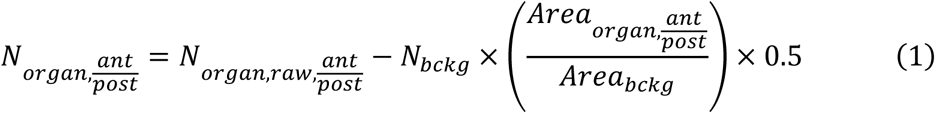

The net organ counts were derived from the anterior and posterior net counts using a conventional geometric mean:

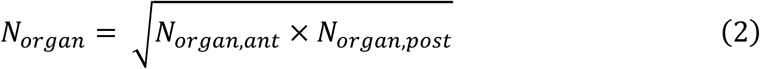

Finally, the LSF was calculated as the ratio of the deposition in the lung to the sum of deposition in the lung and liver:

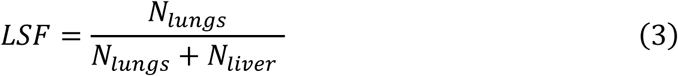

The LSF calculation was done in MIM^TM^ software using the lung shunt fraction workflow. The collection and post-processing of clinical LSF data were approved by the University Institutional Review Board (IRB) as a single-site, retrospective study.

### 3.3 CECT imaging protocol

CECT scans were all performed at UC Davis Health. Twenty-four patients were scanned with a Somatom Definition AS+ (Siemens Healthineers, Erlangen, Germany) system, four patients with a LightSpeed VCT (GE Healthcare, Chicago, IL), one patient with a Sensation 64 (Siemens Healthineers) and one patient with a Revolution Evo (GE Healthcare, Erlangen, Germany). The scan settings included a helical pitch as close as possible to 0.8, a collimation width as close as possible to 40 mm, 100 kV tube voltage, a 0.5-second rotation time, and tube current modulation with an average of 200 mAs. The raw data were reconstructed with 1.0 mm voxels size.

A non-contrast scan was first performed to help identify high-density materials such as calcifications, surgical clips, fiducials, mechanical embolics, and ethiodol related to prior treatments. An iodine-based contrast material with a concentration of 350 mg/mL was delivered via a 30–60 mL bolus injection followed by a 50-mL saline flush. Both the iodine contrast and saline were injected at a rate of 4 mL/sec or higher through an 18– 20-gauge IV cannula.

The late arterial phase, which was captured approximately 40 seconds after the iodine injection, provided the best visualization of highly vascularized tumors, as well as the hepatic arterial tree in which ^90^Y microspheres will be injected and transported by the blood flow ^7,22^.

As the contrast flow rate in the bloodstream varies between patients, the injection-to-scan delay was determined for each patient using a region of interest (ROI) placed on the descending aorta, located in the same axial plane as the liver. The scan was automatically triggered 10 seconds after the ROI average signal density reached a threshold of 160 Hounsfield units (HU). The portal phase was captured 40 seconds after the arterial phase. A delayed phase was taken 3 minutes after injection to image the equilibrium distribution of the contrast agent.

### 3.4 CECT processing

The entire process, including tumor volume estimation, image subtraction, contouring, and LSF estimation is summarized in **Error! Reference source not found.**.

#### 3.4.1 Tumor volume estimation

Reference tumor diameters were estimated retrospectively from the radiology reports to calculate the tumor volumes, assuming a spherical shape. In addition, tumor volumes were contoured slice-by-slice on axial views in the equilibrium phase, allowing for a more accurate estimation of their shape and volume.

Necrotic tumor regions (cyan contours in **Error! Reference source not found.**) were identified as volumes with low enhancement in both the arterial and portal phases. Low-enhancement volumes were contoured independently in each phase using density thresholds calculated by Otsu’s method^33^ applied to the density histogram of each phase (Figure 2). Otsu’s threshold is the local minimum between the two voxel density distributions corresponding to low and normal-to-high enhancement in the liver parenchyma. These low-opacity regions are distinguishable on the histogram as a distinct distribution with reduced opacity compared to the rest of the liver parenchyma, indicative of diminished perfusion. After co-registering both phases using a contour-based hybrid deformable method applied to the whole liver, the contours were intersected. The resulting volume was considered necrotic liver tissue and subtracted from the tumor volume.

#### 3.4.2 Arterial phase subtraction by portal phase

Our hypothesis posits that arteriovenous shunting arises from tumor angiogenesis characterized by unstructured arterial vessels that facilitate shunting. Consequently, tissues with abnormal vascularization exhibit greater opacity in the arterial phase compared to the portal phase and can be visualized by subtracting the portal phase from the late arterial phase and selecting regions where voxels density is near-zero or positive. Other regions not contributing to the LSF are expected to have negative values, as normal, parenchyma with normal vasculature derives blood supply predominantly from the portal system and shows as greater density on the portal and equilibrium phases^34^.

To create the subtraction images, a multistep process is implemented. First, the perfused volume (depicted in red in Figure 1, corresponding to the liver lobe where ^99m^Tc-MAA was injected) was contoured on both the arterial and portal phases. Second, the images are down sampled to 6 mm resolution. Third, the portal phase was subtracted from the arterial phase (Figure 2, left panel). Then, contouring was performed on the image using a density threshold calculated with Otsu’s method^33^ on the perfused volume (**Error! Reference source not found.**, middle panel). The corresponding "positive-to-null" volume is shown in green in Figure 2, right panel. Finally, the necrotic volume, shown in blue in Figure 1, was subtracted from the positive-to-null volume to define the hypervascular volume.

**Figure 1.**
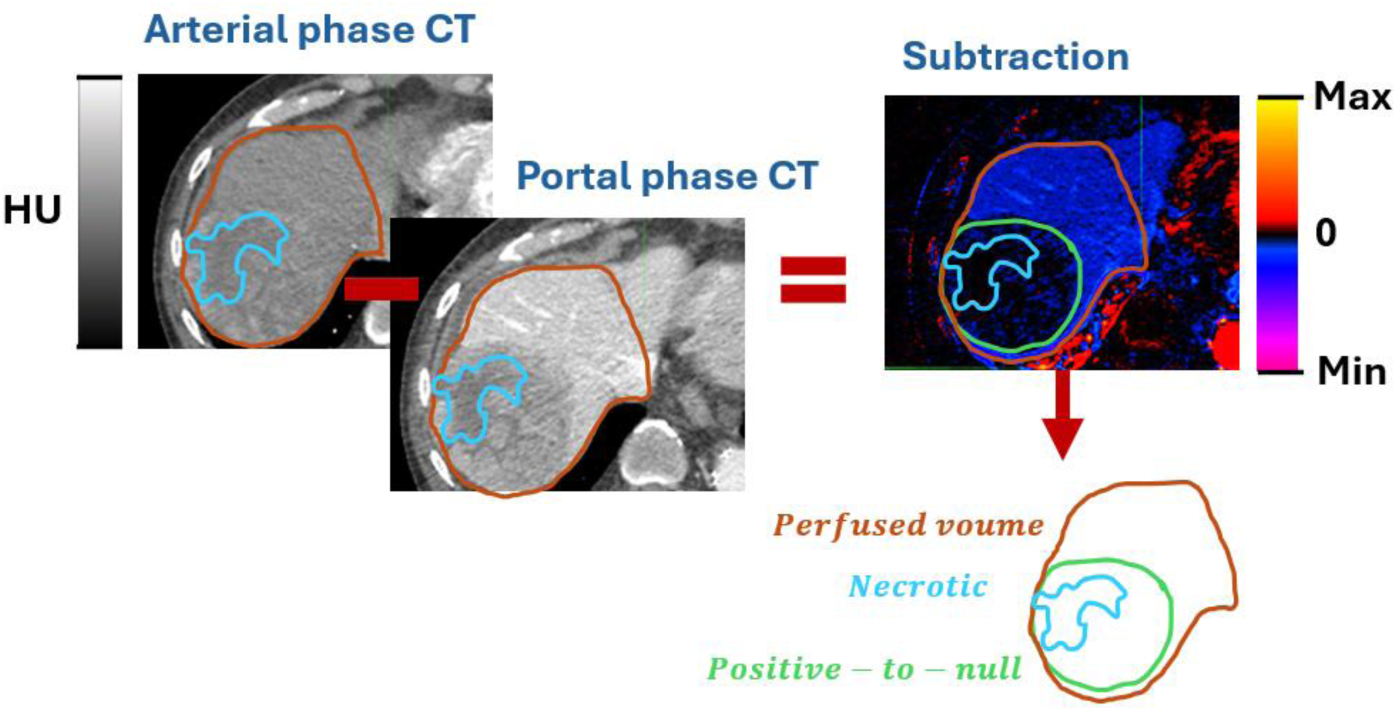
Illustration of perfused, necrotic, and positive-to-null volumes contoured on the CECT late arterial phase, portal phase, and the difference between the portal phase and the arterial phase. The necrotic volume includes regions with low and consistent opacity in both the arterial and portal phases. The perfused volume is typically one lobe, corresponding to the area where Tc-99m MAA is injected. The positive-to-null volume contains voxels with intensity close to or above zero in the subtraction image.

**Figure 2.**
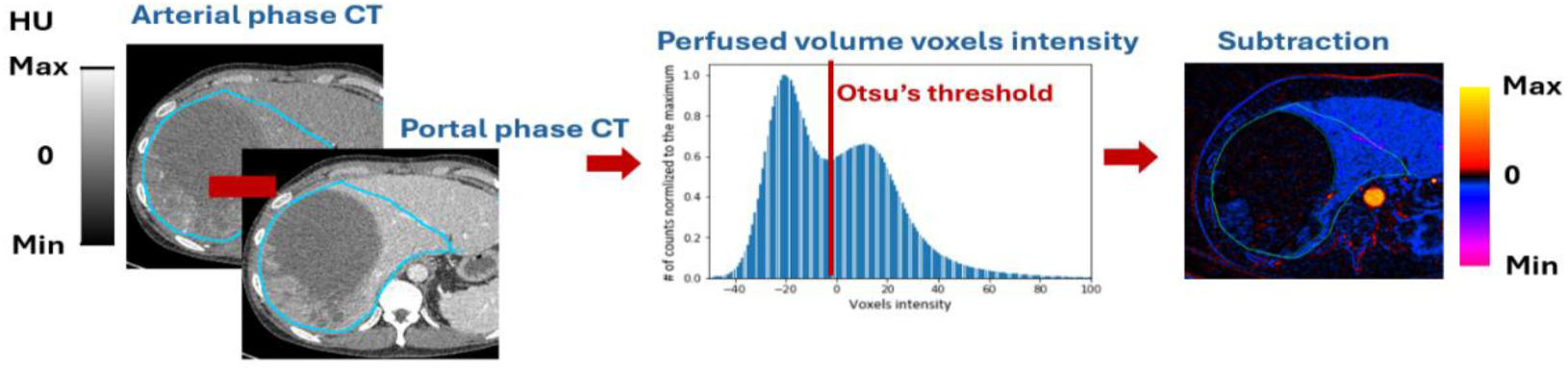
Subtraction of the portal phase from the arterial phase. Otsu’s thresholding is applied to the perfused volume (blue contour, left) on the subtraction image to delineate volumes where voxel intensity is higher or similar in the arterial phase and the portal phase (green contour, right).

### 3.5 Sensitivity analysis

To study the sensitivity of positive-to-null volume and necrotic liver to Otsu’s threshold for contouring, five patients were randomly selected. For each patient, the thresholds derived using Otsu’s method were systematically adjusted by ±5% to evaluate changes in the resulting volumes. This range was chosen to capture realistic variations in thresholding due to noise or image artifacts while maintaining clinical relevance.

### 3.6 Statistical analysis

The study aimed to identify potential predictors of LSF by analyzing various volumetric parameters. Each volume was evaluated by calculating its ratio to the perfused volume. The correlation between these volumes and LSF was assessed using linear regression and the subsequent coefficient of determination (R²). Specifically, R² values were calculated to determine how well each parameter correlated with the LSF measured from planar gamma imaging, considered as ground truth. The evaluated volumes included:

- Tumor volume
- Tumor-to-perfused volume ratio
- Positive-to-null volume
- Positive-to-null-to-perfused volume ratio
- Hypervascular volume
- Hypervascular-to-perfused volume ratio

The Shapiro-Wilk test was utilized to assess the normality of tumor volume distributions, guiding the selection of appropriate statistical methods for further analysis. If the data were normally distributed, a paired t-test was conducted to compare tumor volumes derived from radiologists’ measured diameters and those contoured using the equilibrium phase. Conversely, if tumor volumes deviated from normality, the Wilcoxon Signed-Rank test was employed for this comparison.

## 4 Results

### 4.1 Sensitivity analysis

Results from the sensitivity analysis, presented in Table 1, show a sensitivity to Otsu’s threshold lower or equal to 10% for both positive-to-null volume and necrotic liver determination. This results in an overall impact of less than 10% in the estimation of the positive-to-null-to-perfused volume ratio, hypervascular volume, and hypervascular-to-perfused volume ratio.

**Table 1.** Sensitivity Analysis of Otsu’s Method for Contouring Necrotic Liver and Positive-to-Null Volumes. Threshold variations (±5%) from the calculated Otsu threshold were evaluated to assess their impact on metrics, including hypervascular volume, positive-to-null-to-perfused volume ratio, and hypervascular-to-perfused volume ratio.

|  | Otsu threshold -5% | Otsu threshold +5% |
| --- | --- | --- |
| Necrotic liver tissue | -9% ± 3.1% | 10% ± 3.2% |
| Positive-to-null volume | -6% ± 2.2% | 5% ± 2.1% |
| Positive-to-null-to-perfused volume ratio | -6% ± 2.2% | 5% ± 2.2% |
| Hypervascular volume | -7% ± 3.7% | 8% ± 3.8% |
| Hypervascular-to-perfused volume ratio | -7% ± 3.7% | 8% ± 3.8% |

### 4.2 Relationship between tumor volumes and LSF

The distributions of volumes deduced from diameters reported on the radiology report and of tumor volumes contoured on the equilibrium phase were not normal (p = 2.17×10^-8^ and p = 2.18×10^-8^ for the Shapiro-Wilk test, respectively). No significant difference was found between the two distributions using a Wilcoxon Signed-Rank Test (p = 0.538).

An R² value of 0.34 was obtained from the linear regression between the absolute tumor volumes based on radiology report diameters and the measured LSF, and a R² value of 0.38 when the volumes contoured in the equilibrium phase instead (Figure 3, top row).

**Figure 3.**
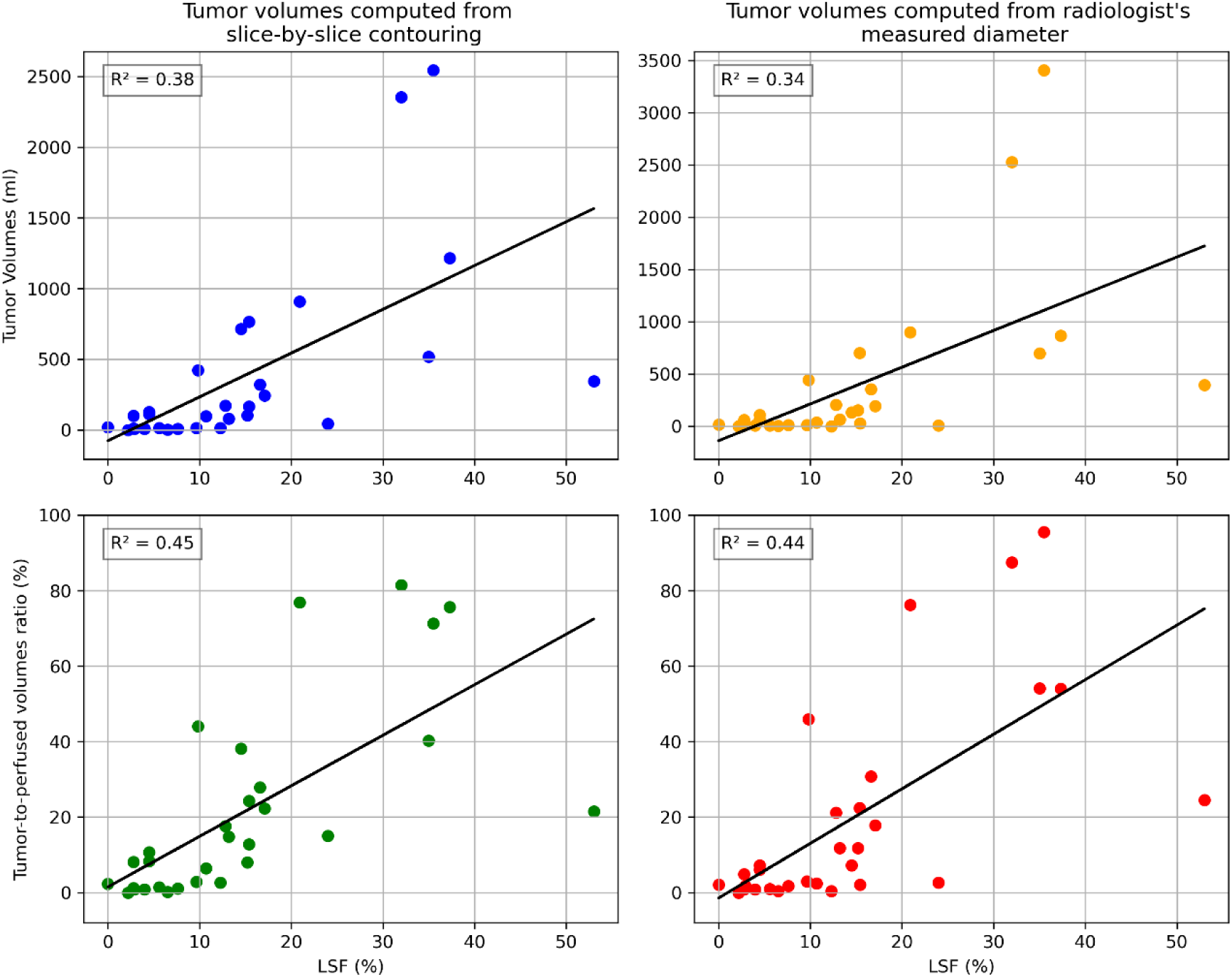
Linear regressions between LSF and tumor volume metrics. Top left: LSF vs. tumor volume from slice-by-slice contouring. Bottom left: LSF vs. ratio between tumor volumes from slice-by-slice contouring and perfused volumes. Top right: LSF vs. tumor volumes from the radiologists’ diameters. Bottom right: LSF vs. ratio between tumor volumes from radiologists’ diameters and perfused volumes. Both tumor volumes computed from radiologists’ measured diameters and tumor volumes from slice-by-slice contouring show similar R² coefficients. R² values increase when the tumor volume is normalized by the perfused volume.

**Figure 4.**
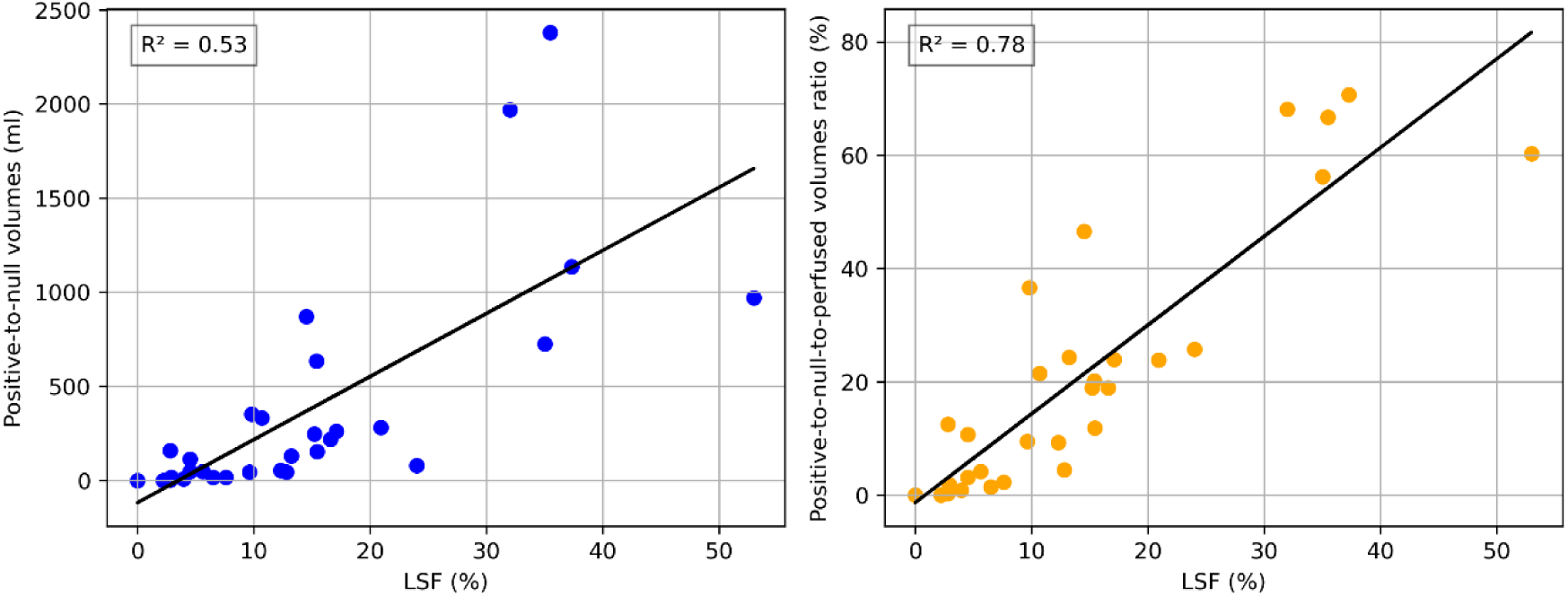
Linear regressions of correlation between LSF and positive-to-null volumes (left) and the ratio positive-to-null-to-perfused volumes ratio (right). The correlation increases when positive-to-null volumes are divided by perfused volumes. The distance between the measured LSF and the regression line also decreases for high LSF values (30%) when normalizing by perfused volumes.

**Figure 5.**
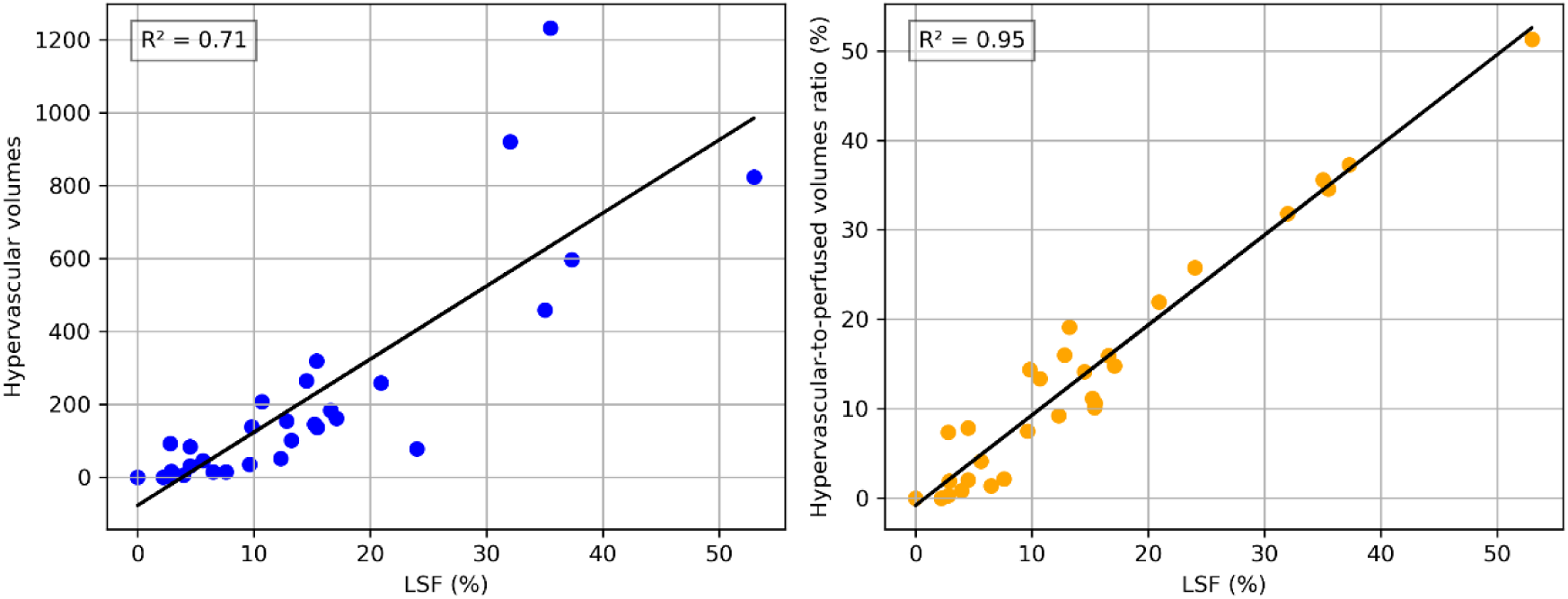
Linear regressions of correlation between LSF and hypervascular volumes (left) and the hypervascular-to-perfused volumes ratio (right). Correlation increases when hypervascular volumes are divided by perfused volumes reaching a R² of 0.95.

An R² value of 0.43 was found from the linear regression between tumor-to-perfused volumes ratio based on radiologists’ measured diameters and the measured LSF from planar gamma imaging, and 0.44 when the volumes contoured in the equilibrium phase instead (Figure 3, bottom row).

### 4.3 Relationship between positive-to-null volumes and LSF

An R² of 0.53 was obtained from the linear regression between the measured positive-to-null volumes and measured LSF. In comparison, the linear regression between positive-to-null-to-perfused volumes and measured LSF gave an R² of 0.78, indicating a stronger correlation (Figure 44).

### 4.4 Hypervascular volumes

An R² of 0.71 was obtained from the linear regression between the hypervascular volumes (as defined in section 3.4.2 as the positive-to-null volume subtracted by the necrotic volume) and measured LSF. In comparison, R² from linear regression between hypervascular-to-perfused volumes ratio and measured LSF was 0.95, indicating a very strong correlation (Figure 45).

The linear regression between the hypervascular-to-perfused volumes ratio and the LSF shows a slope of ∼1 (1.005) and an intercept close to zero (0.008) with standard deviation of 3% for the residuals. The differences between hypervascular-to-perfused volumes ratio and LSF (labeled “residuals” in Figure 66) were found to be normally distributed (p=0.116 with a Shapiro-Wilcoxon test) and suggest that when LSF values increase, the hypervascular-to-perfused volumes ratio becomes a more accurate estimator.

## 5 Discussion

This study investigated the relationship between the LSF measured with standard-of-care ^99m^Tc-MAA planar gamma imaging and volumetric measurements extracted from 4-phase CECT scans, utilizing the differences between arterial and venous phases. This approach aims at understanding if liver perfused territories showing high contrast enhancement in arterial phase correlate with the LSF.

The patient cohort comprises 80% of HCC patients (24 out of 30). The coefficient of correlation between tumor size and LSF was 0.38, matching results from a study with 377 HCC patients (0.36)^28^.

The hypervascular-to-perfused volumes ratio closely correlated with the LSF. Moreover, the predictive power of hypervascular-to-perfused volumes ratio on LSF demonstrates a 1-sigma uncertainty to 3%, suggesting that the hypervascular-to-perfused volumes ratio is directly equal to LSF. The normal distribution of the difference between hypervascular-to-perfused volumes ratio and LSF, closely centered around zero (-0.74 %, Figure 6), indicates the absence of bias. Studies comparing LSF measurements between SPECT/CT and planar imaging found that planar imaging tends to overestimate LSF compared to SPECT/CT with mean values of 8.5% and 4.6%, respectively^14,35^. Additional studies reported on the error in lung mass, LSF, and mean lung dose due to contour variability in patient data (estimating a 1-sigma uncertainty of 13% in lung dose). However, no direct uncertainty estimation of LSF exists in the literature^36^. The 1-sigma uncertainty of 3% obtained with our presented approach is comparable to the discrepancies observed between SPECT/CT and planar imaging methods, indicating its relevance to clinical practice as the errors are comparable to errors accepted in current techniques.

**Figure 6.**
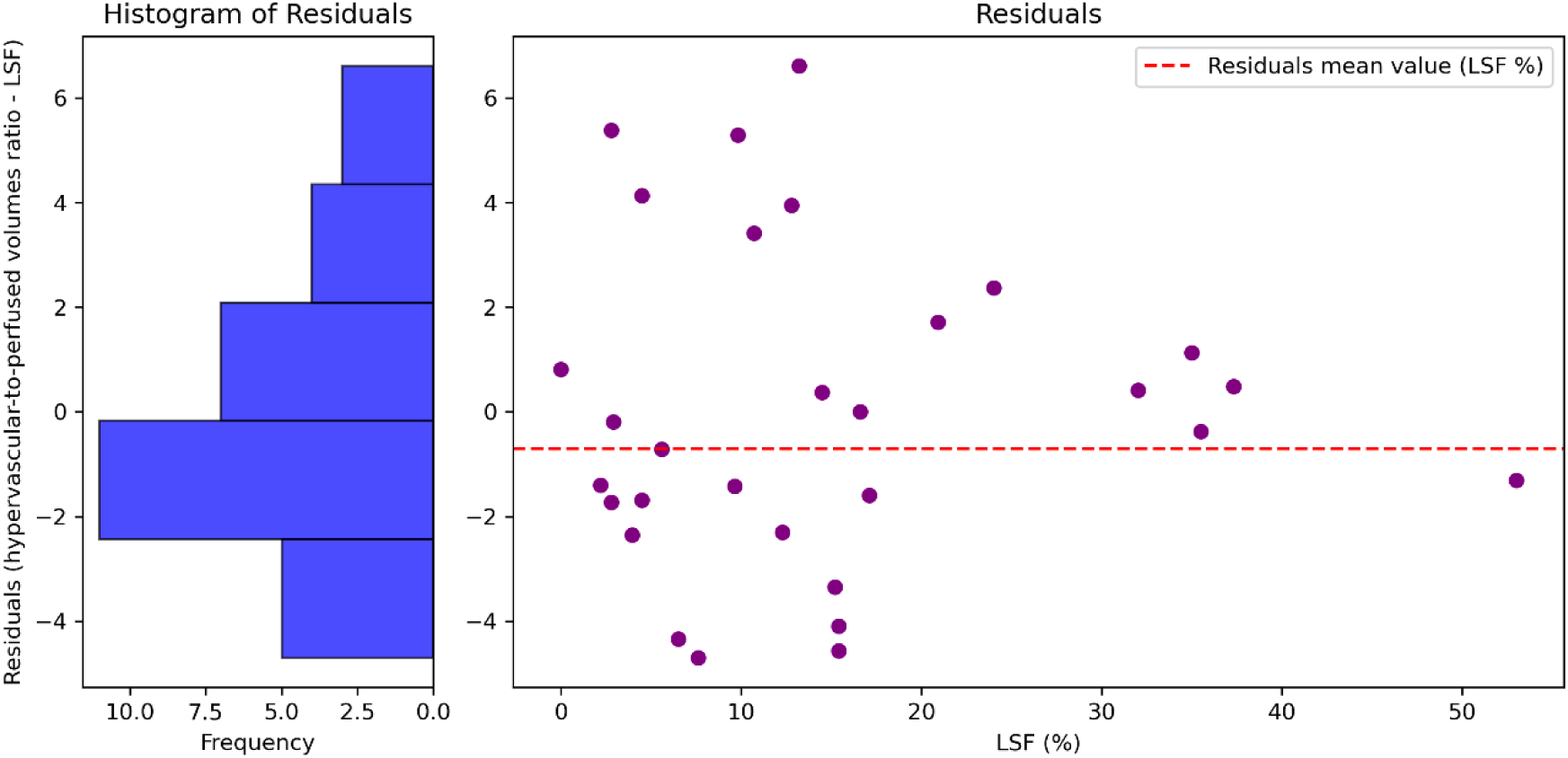
Histogram of differences between hypervascular-to-perfused volumes ratio and LSF (left) and residuals as a function of LSF (right). The mean value of the residuals is -0.71 % LSF (dashed line), suggesting the absence of a systematic error. Residuals also become closer to the average of -0.71% for LSF higher than 20% due to contour errors having less effect on volumes measurements in larger tumors.

The accuracy of LSF estimation appears to improve with increasing hypervascular volumes, likely due to the reduced uncertainty in contoured volume estimations as volumes become larger (>20% of perfused volume; as depicted in Figure 6, where residuals are lower than 2.5%). These findings suggest that residuals and relative residuals decrease with increasing LSF makes the method a potential alternative for high LSF patients since its accuracy improved for LSF greater than 20%. Validation with a larger patient cohort remains necessary to confirm these results.

The strong correlation between hypervascular volumes and LSF, alongside the poor correlation between positive-to-null tissue and LSF, underscores that excluding necrotic tissue from the positive-to-null volumes is crucial for accurate LSF estimation. This can be explained by the absence of blood perfusion in necrotic tissue that cannot exhibit arterio-venous shunting and thus cannot contribute to LSF. This study demonstrates that abnormally high perfusion in the arterial phase, relative to healthy parenchyma and normalized to the perfused volume, serves as a direct indicator of the lung shunt fraction (LSF). In healthy liver parenchyma, the arterial blood supply constitutes only about 25% of the liver’s overall blood supply^34^. By subtracting the portal phase from the arterial phase, areas of abnormally high arterial blood supply become evident with greater contrast in tumoral tissue—a feature directly linked to increased arterio-venous shunting and hence greater LSF^37^.

A patient in the cohort received ^99m^Tc-MAA into a single liver segment and exhibited an LSF of 21% (measured with planar gamma imaging). The hypervascular-to-perfused volume ratio was 20% when considering that segment as the perfused volume but decreased to 5% when considering the whole lobe. This suggests that LSF would have been underestimated if ^99m^Tc-MAA had been administered to the entire lobe. While the literature reports an average LSF of 3.8% in early-stage HCC (UNOS T1/T2/Milan criteria: solitary tumor ≤5 cm or up to 3 nodules each ≤3 cm), these measurements were based on ^99m^Tc-MAA injections into a whole lobe rather than individual segments^19^. No post-treatment lung dosimetry data could be found in the literature to support the increase of LSF in segmentectomy. This highlights the need to better characterize LSF and the clinical impact of ^90^Y deposition in the lung. However, injecting ^99m^Tc-MAA in small volumes is not a suitable strategy because it presents a risk of embolizing small vessels in the tumor vicinity and altering the vascular organization before treatment ^31,38,39^. Our results suggest that CECT could be an alternative to estimate LSF for segmentectomy, without this risk and with superior accuracy.

To enhance the generalizability of our findings, future research will focus on validating these methods across a larger and more diverse patient population, particularly those with metastatic liver cancer. The relatively small number of metastatic patients in the current study may limit the applicability of the results to this group. The retrospective design introduces potential biases, as the study relies on previously collected data that may not fully capture all clinical scenarios. Prospective studies could better assess the reliability of CECT-based LSF estimation in a broader clinical context, removing this selection bias.

Furthermore, a direct comparison with established imaging modalities such as post-treatment SPECT/CT or PET/CT could more directly assess the accuracy of CECT-based LSF estimation. This is also true for standard-of-care radioembolization where the LSF predicted from ^99m^Tc-MAA is rarely verified with post treatment imaging. Variations in CECT imaging protocols across institutions can potentially affect reproducibility, particularly when phase timing differs from those utilized in this study. Standardized imaging protocols may enhance consistency and facilitate wider clinical adoption.

## 6 Conclusion

This study demonstrates that LSF can be estimated using CECT for ^90^Y liver radioembolization, without ^99m^Tc-MAA imaging. The strong correlation between the hypervascular-to-perfused volumes ratio from CECT and LSF from Tc-99m MAA planar imaging suggests that CECT-based methods could serve as a viable alternative to traditional approaches, potentially simplifying treatment planning for Y90 transarterial radioembolization to reduce the risks, costs, and pitfalls of ^99m^Tc-MAA based LSF estimation when using lobal MAA administration to plan selective or superselective Y90 administration. Further validation with larger cohorts will confirm the safety and precision of this approach, which could provide a more efficient and customized method for planning liver radioembolization. Integrating CECT-based LSF estimation into clinical practice could enhance the accuracy and efficiency of ^90^Y radioembolization planning, offering a substantial advancement in the treatment of hepatic malignancies.

## Data Availability

Some data produced in the present study are available upon reasonable request to the authors

